# Predicting Short-Term Mortality in Severe Cirrhosis: An Interpretable Machine Learning Model Integrating Routine Clinical Indicators

**DOI:** 10.1101/2025.07.11.25331343

**Authors:** Shun Zhang, Rui Liu, Zhengjie Li, Tao Pan, Xudong Wen

## Abstract

**Background:** The critical need for precise risk stratification in severe liver cirrhosis is underscored by its substantial 30-day mortality rates, demanding reliable tools to guide clinical interventions.

**Objective:** To establish a machine learning-driven prognostic model for short-term mortality prediction in decompensated cirrhosis through comprehensive analysis of critical care data.

**Methods:** This retrospective cohort study analyzed 1,044 carefully curated cases from the MIMIC-IV database, randomly divided into training (n=740) and validation (n=304) sets. We developed a machine learning model incorporating multidimensional clinical parameters, with rigorous evaluation and internal validation. Short-term survival was analyzed via bootstrap-validated Cox proportional hazards regression. Prognostic heterogeneity across INR-based strata was examined.

**Results:** The final prediction model incorporated eight significant predictors: age (OR 1.040, 95% CI 1.022-1.058), international normalized ratio (OR 1.496, 95%CI 1.297-1726), creatinine (OR 1.210, 95%CI 1.109-1.319), platelets (OR 0.995, 95%CI 0.993-0.997), leukocytes (OR 1.120, 95%CI 1.083-1.159), total bilirubin (OR 1.031, 95%CI 1.006-1.057), peptic ulcer (OR 0.297, 95%CI 0.120-0.734), and metastatic solid tumors (OR 3.001, 95%CI 1.211-7.436). The model demonstrated excellent discrimination with an AUC of 0.851 (95%CI: 0.800–0.901) in the validation cohort. Cox regression analysis confirmed these findings and identified additional associations with aspartate aminotransferase and red blood cell levels. Subgroup analysis revealed significant mortality variations across different INR ranges (P<0.001).

**Conclusions:** Our prediction model identifies high-risk cirrhotic patients and highlights critical prognostic factors, offering clinicians a valuable tool for risk stratification and timely intervention. The strong correlation between laboratory markers, complications, and outcomes underscores the importance of close monitoring in this population.

## 1 Introduction

Liver cirrhosis constitutes a major global health challenge, currently ranking as the 14th leading cause of mortality worldwide. Recent epidemiological data reveal an alarming annual death toll of 1.2 million individuals, with incidence rates projected to escalate through 2039[1, 2]. The clinical trajectory becomes particularly critical at the decompensated stage, which encompasses three distinct clinical phases: stable decompensated cirrhosis, unstable decompensated cirrhosis, and acute-on-chronic liver failure (ACLF). These advanced stages demonstrate disproportionately high mortality rates attributable to complex pathophysiological mechanisms including immune dysregulation, systemic inflammatory responses, and heightened susceptibility to infections[3–5]. Contemporary cohort studies document particularly concerning short-term outcomes: 3-month mortality rates reach 54-58% in decompensated cirrhosis populations[6], while infected cirrhotic patients demonstrate a 63% 1-year mortality rate with a 3.75-fold increased risk compared to non-infected counterparts[7]. The most severe manifestation, ACLF, carries a striking 28-day mortality rate of 66.97%[8]. This escalating mortality burden generates substantial multidimensional impacts, encompassing psychological distress for patients and families, economic strain on healthcare systems, and significant public health challenges. These converging factors underscore the critical imperative for developing precise short-term prognostic models to optimize clinical decision-making and resource allocation.

Despite the availability of multiple prognostic scoring systems, significant methodological limitations hinder their clinical utility in predicting short-term outcomes. The widely adopted MELD/MELD-Na scores, while validated for end-stage survival prediction, demonstrate limited sensitivity in detecting early decompensation events[9, 10]. Similarly, the ALBI score primarily quantifies early-stage hepatic functional decline without capturing multiorgan interactions characteristic of advanced disease[11]. Although nutrition-focused physical assessments like the Subjective Global Assessment show prognostic value for malnutrition-associated mortality, their dependence on clinician expertise introduces interobserver variability[12]. Emerging modalities such as serial liver stiffness measurements exhibit potential in tracking decompensation risk dynamics, yet their isolation from complementary biomarkers restricts comprehensive risk stratification[13]. Notably, frailty indices have demonstrated superior predictive accuracy over MELD-Na for 3-month mortality [14], though implementation barriers persist in routine clinical workflows. This heterogeneity in clinical utility aligns with the multifactorial mortality drivers identified by Vera et al., whose multivariate analysis established hepatic encephalopathy (HR=2.1), infections (HR=3.4), and hepatorenal syndrome (HR=4.7) as independent mortality predictors[15]. These findings collectively underscore the clinical imperative for an integrative prediction framework that synthesizes biochemical, physiological, and complication-specific variables.

This multicenter study pursues three synergistic objectives to address current prognostic limitations in cirrhosis management. First, we propose to design and optimize an interpretable machine learning framework incorporating multifaceted clinical variables: (1) demographic determinants including age, gender, and ethnicity[16]; (2) dynamic laboratory parameters spanning hepatic synthesis markers (bilirubin, albumin), coagulation profiles, and inflammatory indices; (3) extrahepatic comorbidity burdens with emphasis on cardiovascular and renal dysfunction[17, 18]; and (4) validated physiopathological metrics such as MELD-Na, ALBI scores, and objective frailty assessments. Second, through comparative validation across heterogeneous patient cohorts, we aim to quantitatively establish the model’s superior predictive accuracy for 30- and 90-day mortality endpoints relative to conventional scoring systems. Third, as a translational objective, we will develop and deploy an open-access clinical decision support interface with real-time risk stratification capabilities, specifically engineered to facilitate early targeted interventions in high-risk subpopulations.

## 2 Methods

### 2.1 Data Source

We conducted a retrospective cohort study using data from the Medical Information Mart for Intensive Care (MIMIC-IV, version 2.0), a large, freely-available critical care database developed through a collaboration between the Massachusetts Institute of Technology (MIT), Beth Israel Deaconess Medical Center, and Philips Healthcare. After completing the required Collaborative Institutional Training Initiative certification and passing the Protecting Human Research Participants exam, we were granted access to this de-identified database, which waived the requirement for informed consent.We obtained the research data from the MIMIC - IV database on March 8, 2025, and all authors declare that the data we obtained cannot identify the personal characteristics of patients.

### 2.2 Study Design and Participant Selection

Our retrospective analysis utilized hospitalization records from 10,620 patients with cirrhosis diagnoses in the MIMIC-IV critical care database. We implemented rigorous exclusion criteria as detailed in (Supplementary Figure 1): (1) exclusion of non-ICU admissions (n=334), (2) elimination of repeat hospitalizations through retention of only the most recent admission per patient (n=7,140 excluded), and (3) removal of cases with missing critical prognostic variables (n=2,770 excluded). The resultant final analytic cohort consisted of 1,044 unique patients, who underwent stratified randomization via computer-generated allocation sequences to ensure cohort comparability. This process yielded a derivation cohort (n=740) for model development and an independent validation cohort (n=304) for performance evaluation, maintaining a 7:3 ratio consistent with machine learning validation standards (Figure 1).

**Figure 1.**
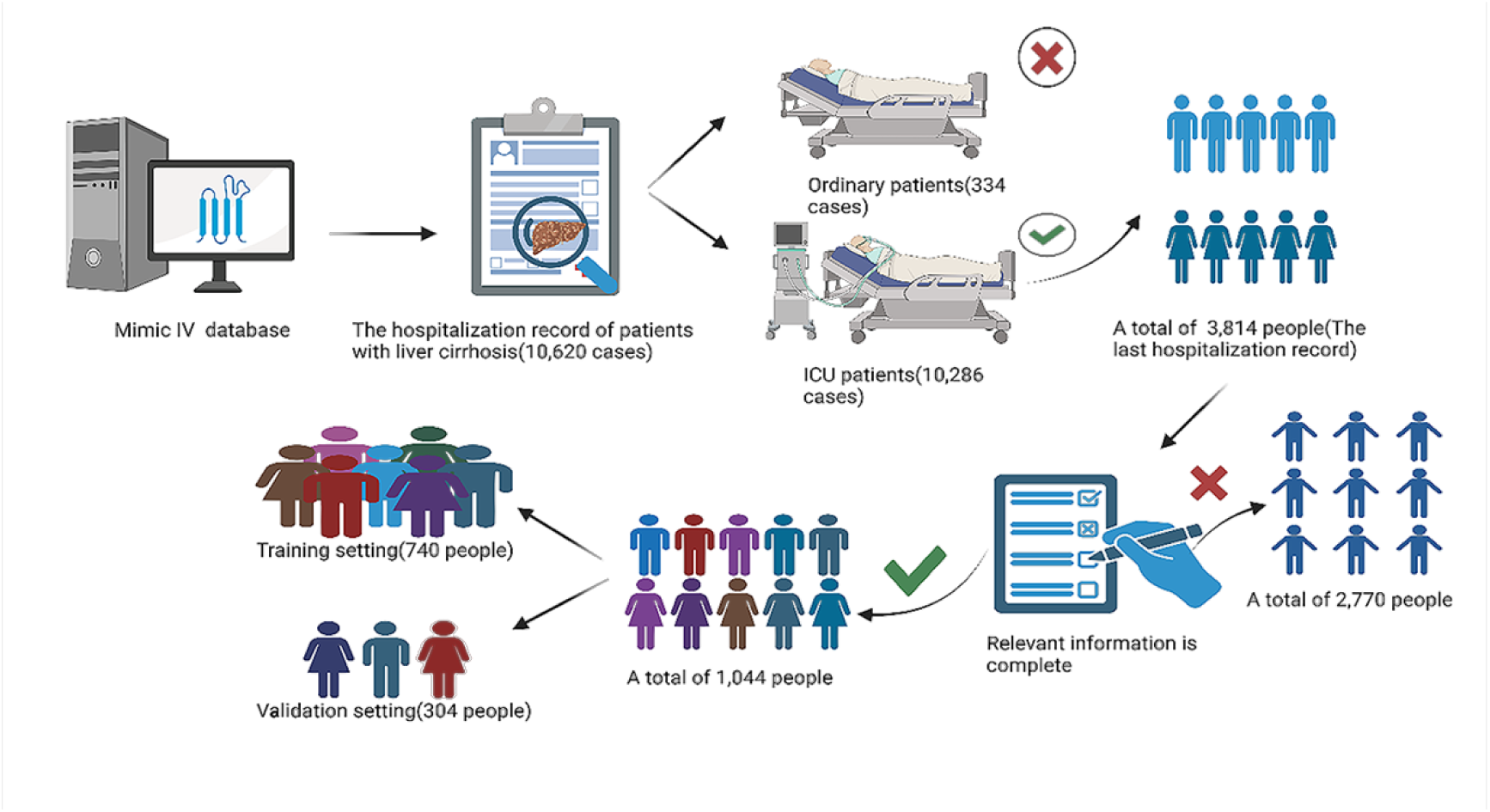
Study cohort selection and data curation workflow.

### 2.3 Data Collection and Processing

Data extraction and harmonization were conducted using Navicat Premium 16 (PremiumSoft CyberTech Ltd.), with comprehensive variable mapping from the MIMIC-IV database. Our structured data collection framework encompassed five core domains: (1) Demographic characteristics including age (years) and admission weight (kg) at ICU entry; (2) Laboratory parameters capturing peak pathological values during ICU stay, comprising hepatic injury profiles (alkaline phosphatase, alanine aminotransferase, aspartate aminotransferase), synthetic function markers (total bilirubin, albumin [minimum recorded value]), hematologic indices (hemoglobin, white blood cell levels, red blood cell levels, platelet counts), renal/metabolic markers (creatinine, blood urea nitrogen, electrolytes [serum sodium minimum, potassium]), and coagulation status (INR); (3) Temporal metrics documenting ICU admission/discharge timelines and mortality endpoints; (4) Validated severity scoring systems (APS III, OASIS, MELD); and (5) Elixhauser-categorized comorbidities with particular attention to cardiovascular (myocardial infarction, congestive heart failure), neurologic (cerebrovascular disease, Alzheimer’s), and systemic disorders (peptic ulcer disease, COPD, diabetes mellitus, renal insufficiency, paralysis, primary/metastatic malignancies). Variables exceeding 20% missingness thresholds were systematically excluded per predefined protocol. All statistical analyses were performed using SPSS (version 29.0; IBM Corp., Armonk, NY), Stata 17 (StataCorp LP, College Station, TX), and R 4.3.1 (R Foundation for Statistical Computing, Vienna, Austria), with X-tile 3.6.2 (Yale University) facilitating optimal cutoff determination through adaptive cohort partitioning.

### 2.4 Primary endpoint

30-day all-cause mortality post-ICU admission.

### 2.5 Statistical Analysis

Continuous variables underwent normality assessment via Kolmogorov-Smirnov tests. Normally distributed variables were expressed as mean ± standard deviation and analyzed using independent samples t-tests with homogeneity of variance verification; non-normally distributed variables were reported as median (interquartile range) and compared with Mann-Whitney U tests. Categorical variables were presented as frequencies (percentages) and analyzed using chi-square tests.A logistic regression model predicting short-term mortality in cirrhotic patients was constructed incorporating age, international normalized ratio (INR), creatinine, platelet count, white blood cell count, total bilirubin, peptic ulcer disease, and metastatic solid tumors. Model performance was evaluated through calibration and discrimination (receiver operating characteristic curve) analyses in the derivation cohort, with subsequent external validation in the independent cohort. A nomogram was developed for visual representation. Decision curve analysis (DCA) quantified clinical utility, while comparative discrimination analyses against MELD and MELD-Na scores demonstrated superior sensitivity and specificity. Internal validation was performed to ensure model robustness.For short-term survival analysis, Cox proportional hazards regression with bootstrap validation (1,000 resamples) was employed. Stratified analyses based on INR thresholds were conducted using Kaplan-Meier methodology to examine mortality incidence across subgroups.

## 3 Results

### 3.1 Baseline Characteristics

The study included 1,044 patients divided into derivation (n=740) and validation cohorts (n=304). Baseline characteristics were generally balanced between cohorts (*P*>0.05 for all comparisons), except for marginally higher prothrombin time international normalized ratio (INR) and renal disease prevalence in the derivation cohort; however, randomization minimized selection bias, and these differences were deemed clinically non-significant.

In the derivation cohort, 204 deaths (27.6%) occurred within 30 days. Compared with survivors, deceased patients exhibited significantly higher age, weight, liver enzymes (ALT, ALP, AST), INR, creatinine, white blood cell count (WBC), blood urea nitrogen (BUN), and total bilirubin, alongside lower hemoglobin, albumin, red blood cell count (RBC), sodium, and platelet levels (all *P*<0.05). Comorbidities including myocardial infarction, congestive heart failure, cerebrovascular disease, chronic pulmonary disease, renal disease, malignancy, liver disease, and metastatic solid tumors were more prevalent in the death group (*P*<0.05).

Univariate logistic regression identified 12 mortality predictors (*P*<0.05): age, albumin, sodium, RBC, ALP, AST, INR, creatinine, platelets, WBC, BUN, total bilirubin (clinical variables), and cerebrovascular disease, peptic ulcer disease, renal disease, and metastatic solid tumors (comorbidities). Complete baseline comparisons and univariate results are detailed in (Tables 1 and Table 2), respectively.

**Table1.** Characteristics at baseline of patients in the study.

|  | Training Setting | Validation Setting | P |
| --- | --- | --- | --- |
| Variable | (n=740) | (n=304) | Value |
| Age (years) | 60.24±12.22 | 59.54±12.16 | 0.403 |
| Weight (kg) | 84.99±21.48 | 85.25±20.32 | 0.856 |
| Hemoglobin (max, g/dl) | 10.21±1.96 | 10.04±1.88 | 0.192 |
| Albumin (min, g/dl) | 2.71±0.56 | 2.69±0.55 | 0.650 |
| RBC (max, 10<sup>6</sup>/ul) | 3.26±0.67 | 3.21±0.68 | 0.299 |
| Sodium (min, mmol/l) | 132.28±6.12 | 132.08±5.57 | 0.614 |
| ALT (max, u/l) | 58.00 (30.00-153.00) | 61.00 (30.25-121.25) | 0.480 |
| ALP(max, u/l) | 110.00 (74.00-152.47) | 115.00 (77.00-155.00) | 0.278 |
| AST(max, u/l) | 112.50 (57.00-289.50) | 104.5 (51.00-219.25) | 0.331 |
| INR(max) | 1.80 (1.40-2.70) | 2.00 (1.50-3.18) | 0.038 |
| Creatinine (max, mg/dl) | 1.60 (0.90-3.00) | 1.60 (1.00-3.00) | 0.625 |
|  | 150.00 (102.00- |  |  |
| Platelet (max, 10<sup>9</sup>/l) | 240.75) | 157.00 (101.25-234.50) | 0.758 |
| WBC (max, 10<sup>9</sup>/l) | 9.30 (7.73-9.80) | 9.40 (7.60-9.80) | 0.507 |
| BUN(max, mg/dl) | 37.00 (22.00-65.00) | 39.50 (21.00-69.50) | 0.708 |
| Bilirubin total (max, mg/dl) | 3.60 (1.50-8.40) | 3.80 (1.80-9.08) | 0.143 |
| **Comorbidity** |  |  |  |
| Myocardial infarct |  |  |  |
| Yes | 42 (5.7%) | 20 (6.6%) | 0.575 |
| No | 698 (94.3%) | 284 (93.4%) |  |
| Congestive heart failure |  |  |  |
| Yes | 146 (19.7%) | 49 (16.1%) | 0.174 |
| No | 594 (80.3%) | 255 (83.9%) |  |
| Peripheral vascular disease |  |  |  |
| Yes | 50 (6.8%) | 30 (9.9%) | 0.086 |
| No | 690 (93.2%) | 274 (90.1%) |  |
| Cerebrovascular disease |  |  |  |
| Yes | 54 (7.3%) | 25 (8.2%) | 0.607 |
| No | 686 (92.7%) | 279 (91.8%) |  |
| Chronic pulmonary disease |  |  |  |
| Yes | 181 (24.5%) | 75 (24.7%) | 0.942 |
| No | 559 (75.5%) | 229 (75.3%) |  |
| Peptic ulcer disease |  |  |  |
| Yes | 56 (7.6%) | 15 (4.9%) | 0.125 |
| No | 684 (92.4%) | 289 (95.1%) |  |
| Renal disease |  |  |  |
| Yes | 172 (23.2%) | 53 (17.4%) | 0.038 |
| No | 568 (76.8%) | 251 (82.6%) |  |
| Malignant cancer |  |  |  |
| Yes | 116 (15.7%) | 49 (16.1%) | 0.859 |
| No | 624 (84.3%) | 255 (83.9%) |  |
| Severe liver disease |  |  |  |
| Yes | 430 (58.1%) | 181 (59.5%) | 0.670 |
| No | 310 (41.9%) | 123 (40.5%) |  |
| Metastatic solid tumor |  |  |  |
| Yes | 29 (3.9%) | 14 (4.6%) | 0.612 |
| No | 711 (96.1%) | 290 (95.4%) |  |

**Table2.**
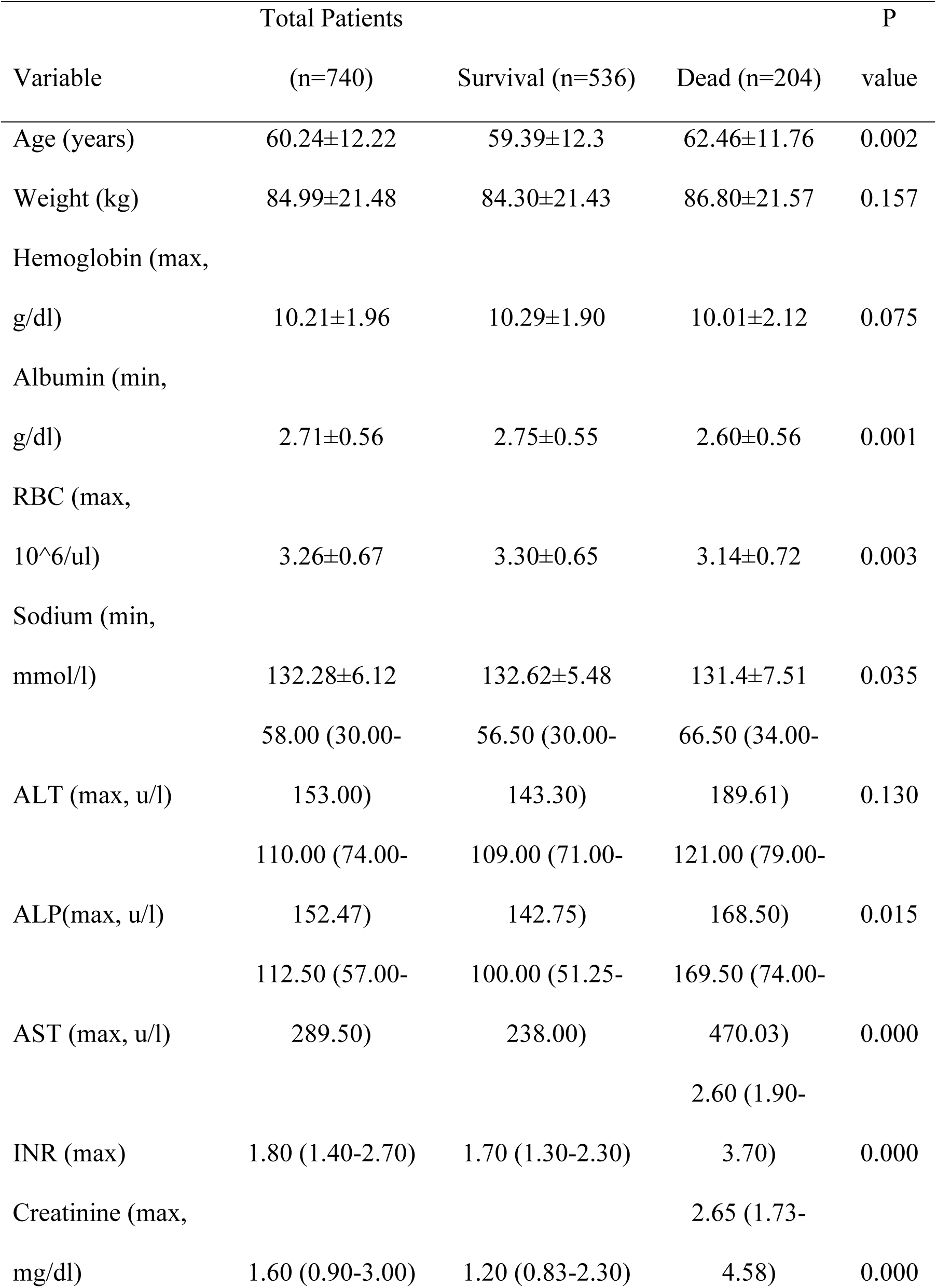
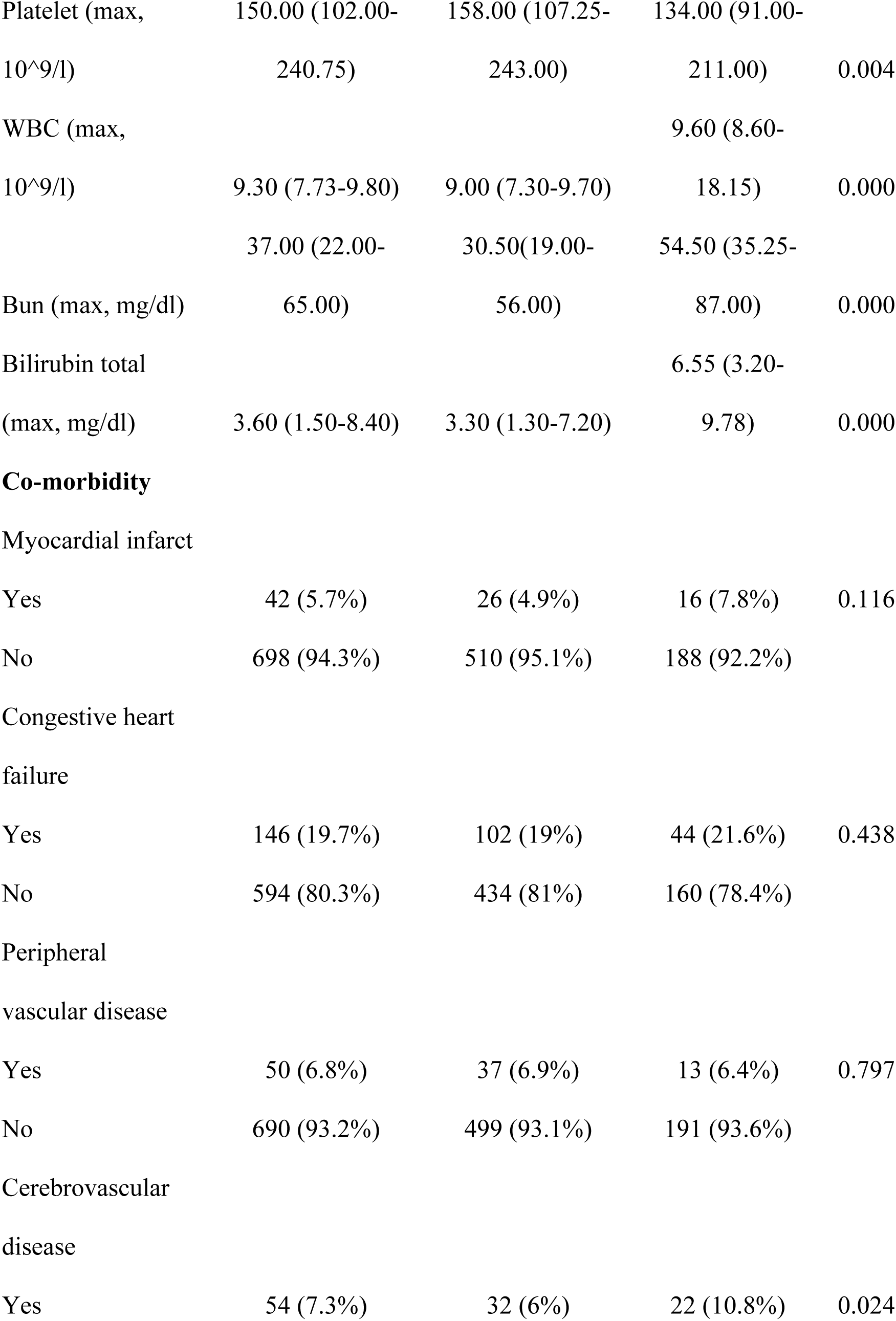
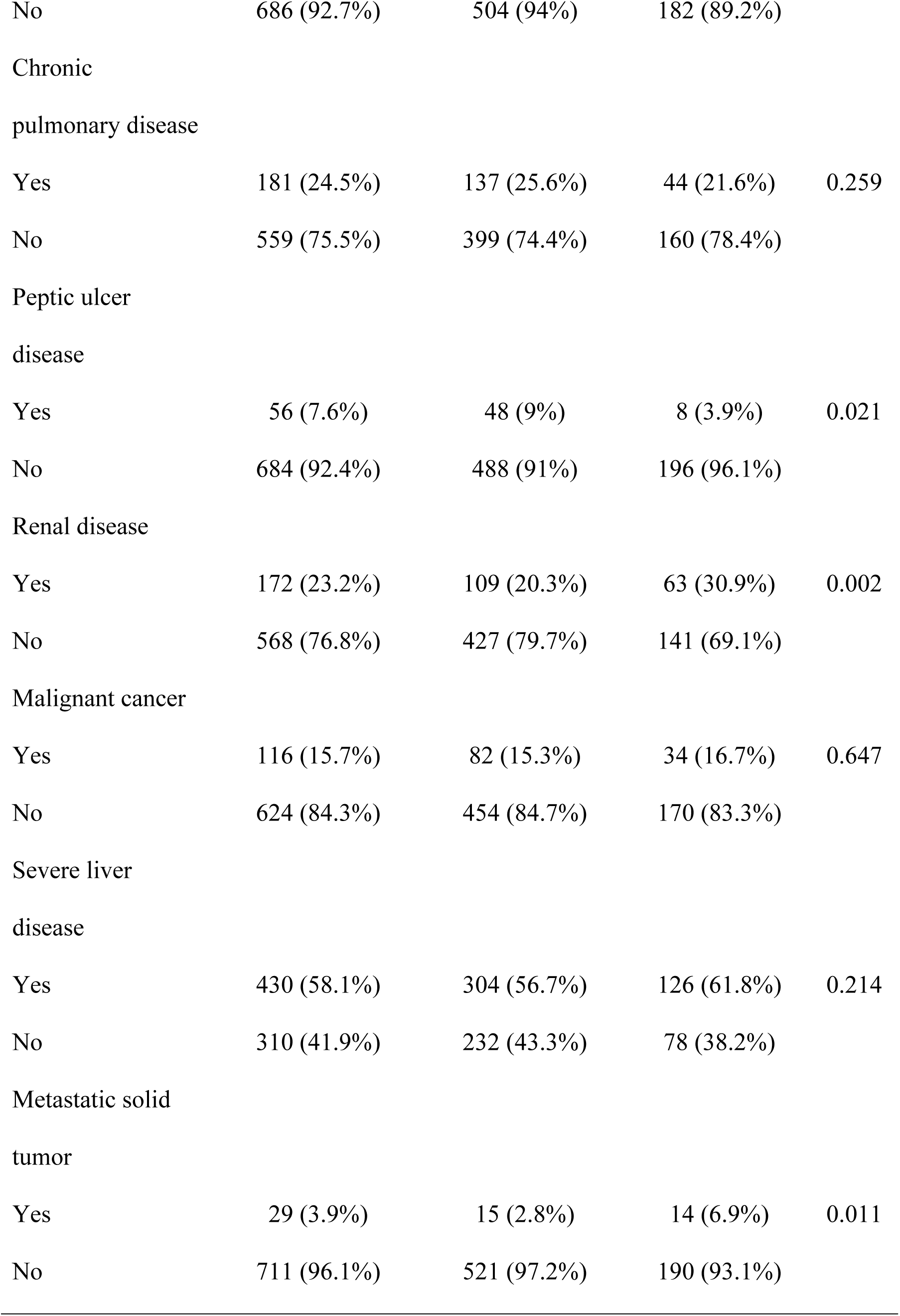
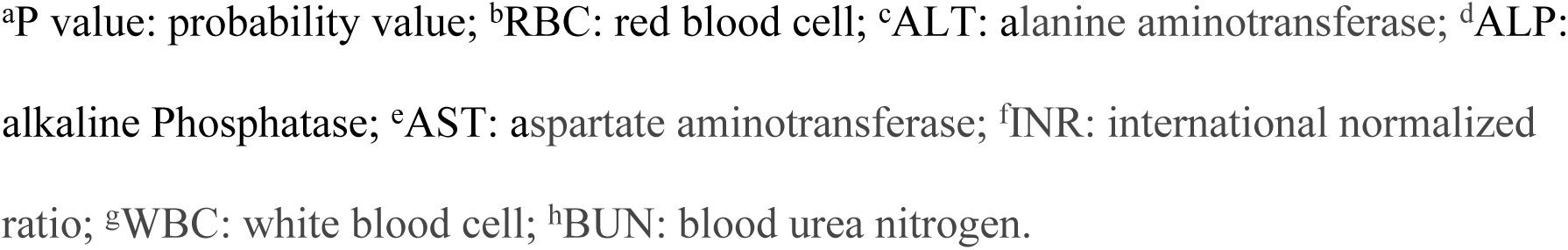
Baseline Analysis of the training cohort.

| Variable | Total Patients |  |  | P |
| --- | --- | --- | --- | --- |
|  | (n=740) | Survival (n=536) | Dead (n=204) | value |
| Age (years) | 60.24±12.22 | 59.39±12.3 | 62.46±11.76 | 0.002 |
| Weight (kg) | 84.99±21.48 | 84.30±21.43 | 86.80±21.57 | 0.157 |
| Hemoglobin (max,<br>g/dl) | 10.21±1.96 | 10.29±1.90 | 10.01±2.12 | 0.075 |
| Albumin (min,<br>g/dl) | 2.71±0.56 | 2.75±0.55 | 2.60±0.56 | 0.001 |
| RBC (max,<br>10<sup>6</sup>/ul) | 3.26±0.67 | 3.30±0.65 | 3.14±0.72 | 0.003 |
| Sodium (min,<br>mmol/l) | 132.28±6.12 | 132.62±5.48 | 131.4±7.51 | 0.035 |
| ALT (max, u/l) | 58.00 (30.00- | 56.50 (30.00- | 66.50 (34.00- | 0.130 |
|  | 153.00) | 143.30) | 189.61) |  |
| ALP(max, u/l) | 110.00 (74.00- | 109.00 (71.00- | 121.00 (79.00- | 0.015 |
|  | 152.47) | 142.75) | 168.50) |  |
| AST (max, u/l) | 112.50 (57.00- | 100.00 (51.25- | 169.50 (74.00- | 0.000 |
|  | 289.50) | 238.00) | 470.03) |  |
| INR (max) | 1.80 (1.40-2.70) | 1.70 (1.30-2.30) | 2.60 (1.90- | 0.000 |
|  |  |  | 3.70) |  |
| Creatinine (max,<br>mg/dl) | 1.60 (0.90-3.00) | 1.20 (0.83-2.30) | 2.65 (1.73- | 0.000 |
|  |  |  | 4.58) |  |
| Platelet (max,<br>10<sup>9</sup>/l) | 150.00 (102.00-240.75) | 158.00 (107.25-243.00) | 134.00 (91.00-211.00) | 0.004 |
| WBC (max,<br>10<sup>9</sup>/l) | 9.30 (7.73-9.80) | 9.00 (7.30-9.70) | 9.60 (8.60-18.15) | 0.000 |
| Bun (max, mg/dl) | 37.00 (22.00-65.00) | 30.50(19.00-56.00) | 54.50 (35.25-87.00) | 0.000 |
| Bilirubin total<br>(max, mg/dl) | 3.60 (1.50-8.40) | 3.30 (1.30-7.20) | 6.55 (3.20-9.78) | 0.000 |
| **Co-morbidity** |  |  |  |  |
| Myocardial infarct |  |  |  |  |
| Yes | 42 (5.7%) | 26 (4.9%) | 16 (7.8%) | 0.116 |
| No | 698 (94.3%) | 510 (95.1%) | 188 (92.2%) |  |
| Congestive heart<br>failure |  |  |  |  |
| Yes | 146 (19.7%) | 102 (19%) | 44 (21.6%) | 0.438 |
| No | 594 (80.3%) | 434 (81%) | 160 (78.4%) |  |
| Peripheral<br>vascular disease |  |  |  |  |
| Yes | 50 (6.8%) | 37 (6.9%) | 13 (6.4%) | 0.797 |
| No | 690 (93.2%) | 499 (93.1%) | 191 (93.6%) |  |
| Cerebrovascular<br>disease |  |  |  |  |
| Yes | 54 (7.3%) | 32 (6%) | 22 (10.8%) | 0.024 |
| No | 686 (92.7%) | 504 (94%) | 182 (89.2%) |  |
| Chronic<br>pulmonary disease |  |  |  |  |
| Yes | 181 (24.5%) | 137 (25.6%) | 44 (21.6%) | 0.259 |
| No | 559 (75.5%) | 399 (74.4%) | 160 (78.4%) |  |
| Peptic ulcer<br>disease |  |  |  |  |
| Yes | 56 (7.6%) | 48 (9%) | 8 (3.9%) | 0.021 |
| No | 684 (92.4%) | 488 (91%) | 196 (96.1%) |  |
| Renal disease |  |  |  |  |
| Yes | 172 (23.2%) | 109 (20.3%) | 63 (30.9%) | 0.002 |
| No | 568 (76.8%) | 427 (79.7%) | 141 (69.1%) |  |
| Malignant cancer |  |  |  |  |
| Yes | 116 (15.7%) | 82 (15.3%) | 34 (16.7%) | 0.647 |
| No | 624 (84.3%) | 454 (84.7%) | 170 (83.3%) |  |
| Severe liver<br>disease |  |  |  |  |
| Yes | 430 (58.1%) | 304 (56.7%) | 126 (61.8%) | 0.214 |
| No | 310 (41.9%) | 232 (43.3%) | 78 (38.2%) |  |
| Metastatic solid<br>tumor |  |  |  |  |
| Yes | 29 (3.9%) | 15 (2.8%) | 14 (6.9%) | 0.011 |
| No | 711 (96.1%) | 521 (97.2%) | 190 (93.1%) |  |

<sup>a</sup>P value: probability value; <sup>b</sup>RBC: red blood cell; <sup>c</sup>ALT: alanine aminotransferase; <sup>d</sup>ALP: alkaline Phosphatase; <sup>e</sup>AST: aspartate aminotransferase; <sup>f</sup>INR: international normalized ratio; <sup>g</sup>WBC: white blood cell; <sup>h</sup>BUN: blood urea nitrogen.

### 3.2 Model Development

Multivariable logistic regression identified eight independent predictors of 30-day mortality in severe cirrhosis (Table 3; visualized via nomogram in Figure 2A): age (adjusted odds ratio [aOR] 1.040, 95% CI 1.022–1.058 per year), international normalized ratio (INR; aOR 1.496, 95% CI 1.297–1.726 per unit), creatinine (aOR 1.210, 95% CI 1.109–1.319 mg/dL), platelets (aOR 0.995, 95% CI 0.993–0.997 ×10³/μL), leukocytes (aOR 1.120, 95% CI 1.083–1.159 ×10³/μL), total bilirubin (aOR 1.031, 95% CI 1.006–1.057 mg/dL), peptic ulcer disease (aOR 0.297, 95% CI 0.120–0.734), and metastatic solid tumors (aOR 3.001, 95% CI 1.211–7.436). Key clinical interpretations revealed: (1) 4% incremental mortality risk per year of age and 50% per INR unit increase (P<0.001 for both); (2) metastatic solid tumors tripled mortality risk (*P*<0.001); (3) peptic ulcer disease paradoxically reduced risk by 70% (aOR 0.297; potential confounding addressed in Limitations). The nomogram achieved clinically actionable stratification (Figure 2B), with low-risk (<5 points) and high-risk (>15 points) groups demonstrating <0.1% and 99% predicted mortality, respectively.

**Table3.** Univariate and multivariate Logistic regression analyses.

| Variables | Univariate logistic model | Multivariable logistic model |
| --- | --- | --- |

|  | OR | 95%CI | P-value | OR | 95%CI | P-Value |
| --- | --- | --- | --- | --- | --- | --- |
|  | 1.02 | 1.007- | 0.00 | 1.04 | 1.022- |  |
| Age | 1 | 1.035 | 2 | 0 | 1.058 | 0.000 |
|  | 1.00 | 1.001- | 0.00 |  |  |  |
| ALP | 2 | 1.004 | 4 |  |  |  |
|  | 1.00 | 1.000- | 0.00 |  |  |  |
| AST | 0 | 1.000 | 0 |  |  |  |
|  | 1.65 | 1.452- | 0.00 | 1.49 | 1.297- |  |
| INR | 3 | 1.883 | 0 | 6 | 1.726 | 0.000 |
|  | 1.26 | 1.171- | 0.00 | 1.31 | 1.109- |  |
| Creatinine | 0 | 1.356 | 0 | 9 | 1.319 | 0.000 |
|  | 0.99 | 0.997- | 0.01 | 0.99 | 0.993- |  |
| Platelet | 8 | 1.000 | 9 | 5 | 0.997 | 0.000 |
|  | 0.68 | 0.532- | 0.00 |  |  |  |
| RBC | 6 | 0.885 | 4 |  |  |  |
|  | 1.11 | 1.078- | 0.00 | 1.12 | 1.083- |  |
| WBC | 0 | 1.143 | 0 | 0 | 1.059 | 0.000 |
|  | 0.61 | 0.453- | 0.00 |  |  |  |
| Albumin | 0 | 0.823 | 1 |  |  |  |
|  | 0.96 | 0.944- | 0.01 |  |  |  |
| Sodium | 9 | 0.994 | 6 |  |  |  |
|  | 1.01 | 1.013- | 0.00 |  |  |  |
| BUN | 8 | 1.023 | 0 |  |  |  |
|  | 1.05 | 1.041- | 0.00 | 1.03 | 1.006- |  |
| Bilirubin total | 2 | 1.083 | 0 | 1 | 1.057 | 0.004 |
| Cerebrovascular | 1.90 | 1.078- | 0.02 |  |  |  |
| disease | 4 | 3.362 | 6 |  |  |  |
|  | 0.41 | 0.193- | 0.02 | 0.29 | 0.120- |  |
| Peptic ulcer disease | 5 | 0.893 | 5 | 7 | 0.734 | 0.009 |
|  | 1.75 | 1.216- | 0.00 |  |  |  |
| Renal disease | 0 | 2.519 | 3 |  |  |  |
|  | 2.55 | 1.213- | 0.01 | 3.00 | 1.211- | 0.01 |
| Metastatic solid tumor | 9 | 5.402 | 4 | 1 | 7.436 | 8 |

<sup>a</sup>P value:probability value; <sup>b</sup>RBC: red blood cell; <sup>c</sup>ALT: alanine aminotransferase; <sup>d</sup>ALP: alkaline Phosphatase; <sup>e</sup>AST: aspartate aminotransferase; <sup>f</sup>INR: international normalized ratio; <sup>g</sup>WBC: white blood cell; <sup>h</sup>BUN: blood urea nitrogen

**Figure 2.**
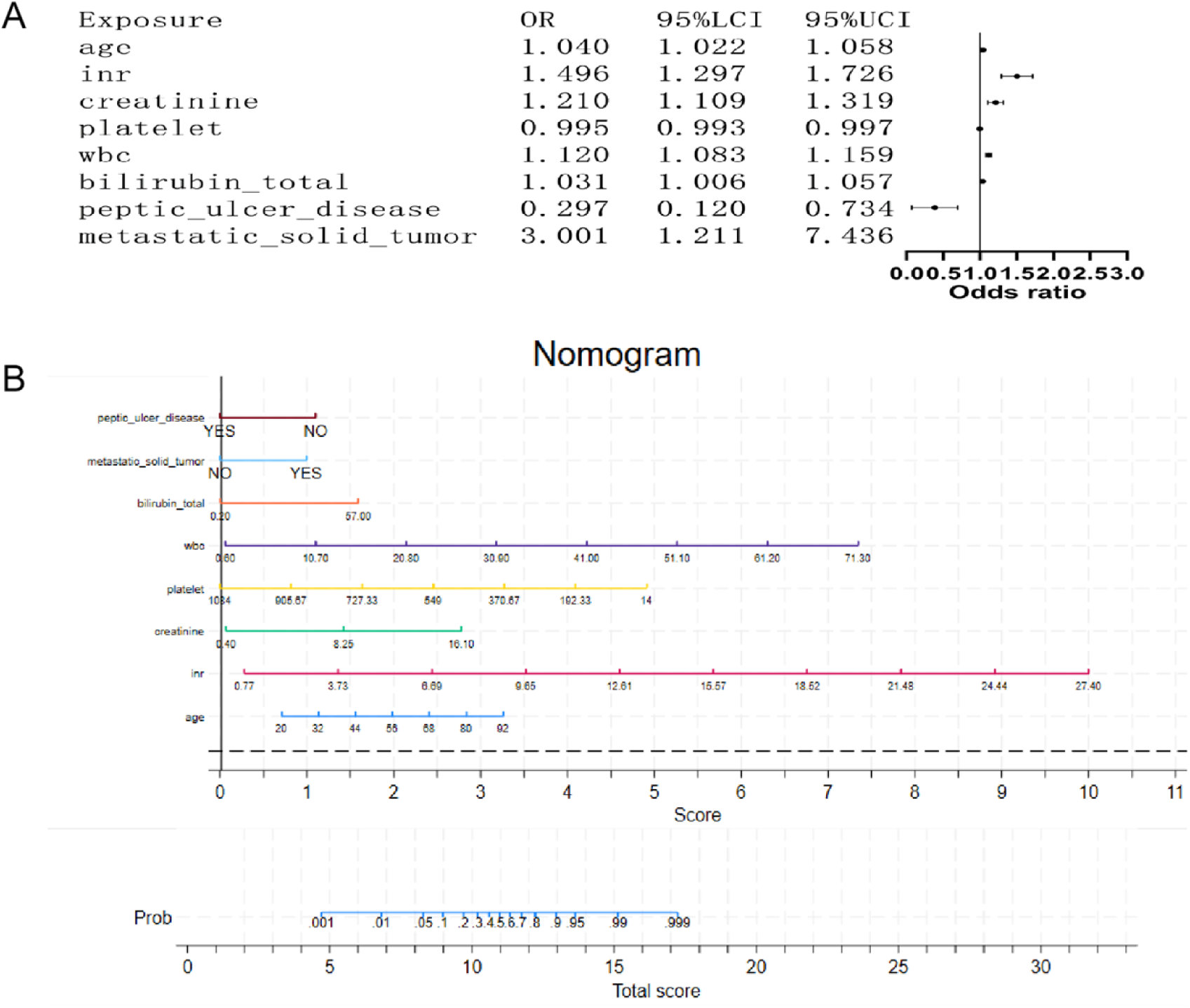
Multivariable analysis and nomogram development for short-term mortality prediction in severe cirrhosis. (A) Forest plot of adjusted odds ratios (aORs) derived from multivariable logistic regression. Key predictors include age (aOR 1.040 per year, 95% CI: 1.022–1.058), INR (aOR 1.496, 95% CI: 1.297–726), and creatinine (aOR 1.210, 95% CI: 1.109–1.319). All covariates met proportional hazards assumptions (p< 0.05). (B) Clinically actionable nomogram integrating significant predictors to estimate individualized 30-day mortality probabilities. Points are assigned per variable value, with total scores mapped to predicted risk (range: 0.1%–99.9%).

### 3.3 Model Performance Evaluation

#### Model Calibration

The nomogram demonstrated high calibration accuracy for 30-day mortality prediction in severe cirrhosis across both derivation and validation cohorts. Quantitative assessments revealed: (1) observed-to-expected mortality ratio of 1.00, indicating near-perfect agreement between predicted and actual outcomes; (2) calibration-in-the-large intercept of 0 (95% CI -0.12 to 0.11), confirming absence of systematic over- or under-prediction bias. Visual calibration plots further validated model precision, with predicted probabilities closely aligned along the 45° ideal reference line in both cohorts (Figures 3A and 3B).

**Figure 3.**
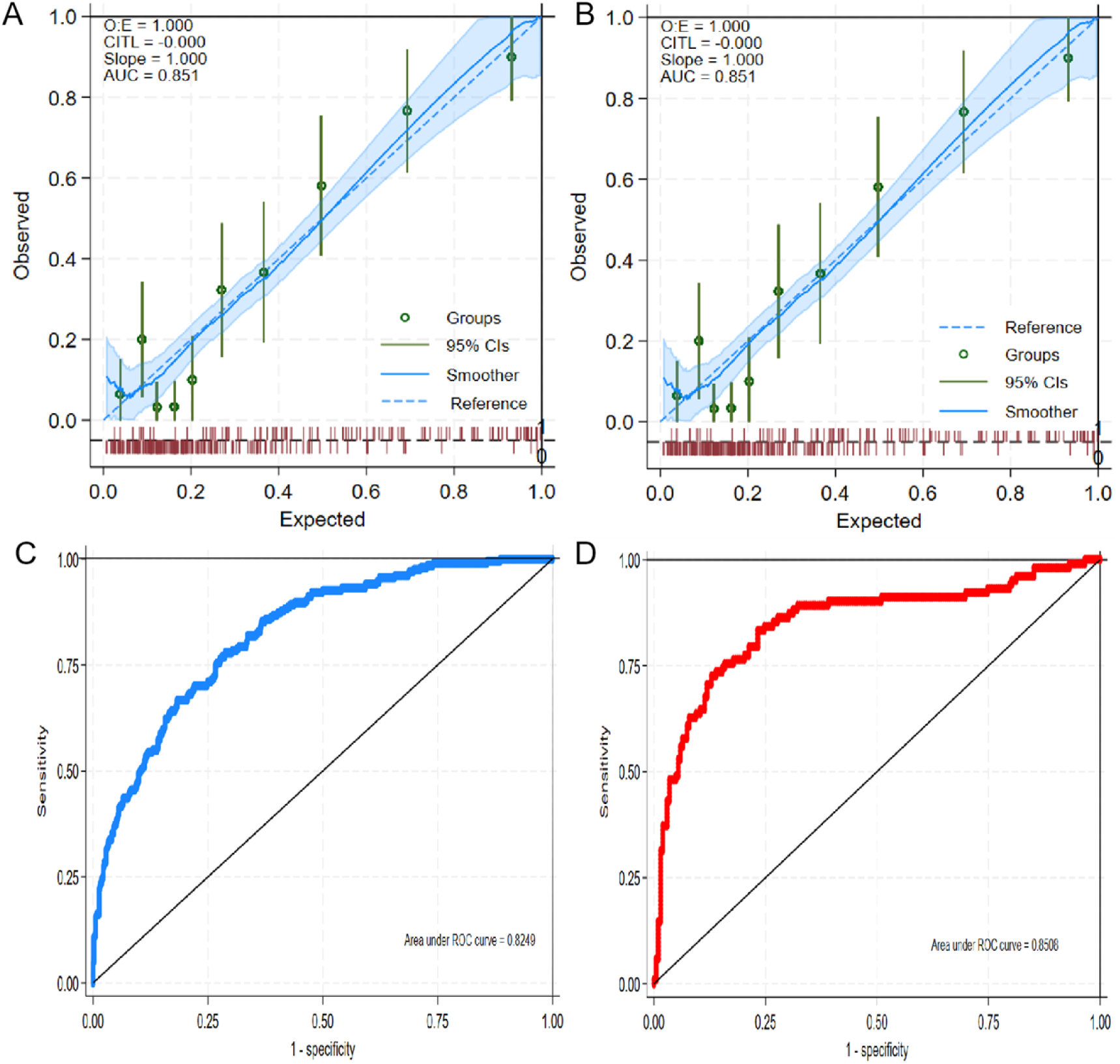
Calibration and discrimination performance of the short-term mortality prediction model in severe cirrhosis. (A) Calibration curve for the derivation cohort, demonstrating agreement between predicted mortality probabilities (x-axis) and observed outcomes (y-axis). The dashed diagonal represents perfect calibration, while the solid blue line indicates model performance (Brier score <0.25; Hosmer-Lemeshow test p>0.05). (B) Validation cohort calibration curve showing preserved accuracy across risk strata (Brier score <0.25; p>0.05). (C) Receiver operating characteristic (ROC) curve for the derivation cohort, with the proposed model achieving an AUC of 0.825 (95% CI: 0.793–0.857). (D) External validation ROC curve confirming robust discrimination AUC = 0.851 (95% CI: 0.800– 0.901).

#### Discrimination Performance

Receiver operating characteristic (ROC) curve analysis demonstrated robust discriminative capacity, with area under the curve (AUC) of 0.825 (95% CI 0.793–0.857) in the derivation cohort and 0.851 (95% CI 0.800–0.901) in the validation cohort (Figures 3C and 3D). When benchmarked against conventional scoring systems, our model exhibited superior discriminative ability: derivation cohort AUC improved by 0.088–0.090 compared to MELD (AUC 0.737) and MELD-Na (AUC 0.741), while validation cohort AUC exceeded both scores by 0.059–0.067 (MELD: 0.784; MELD-Na: 0.792) (Figures 4A and 4B).

**Figure 4.**
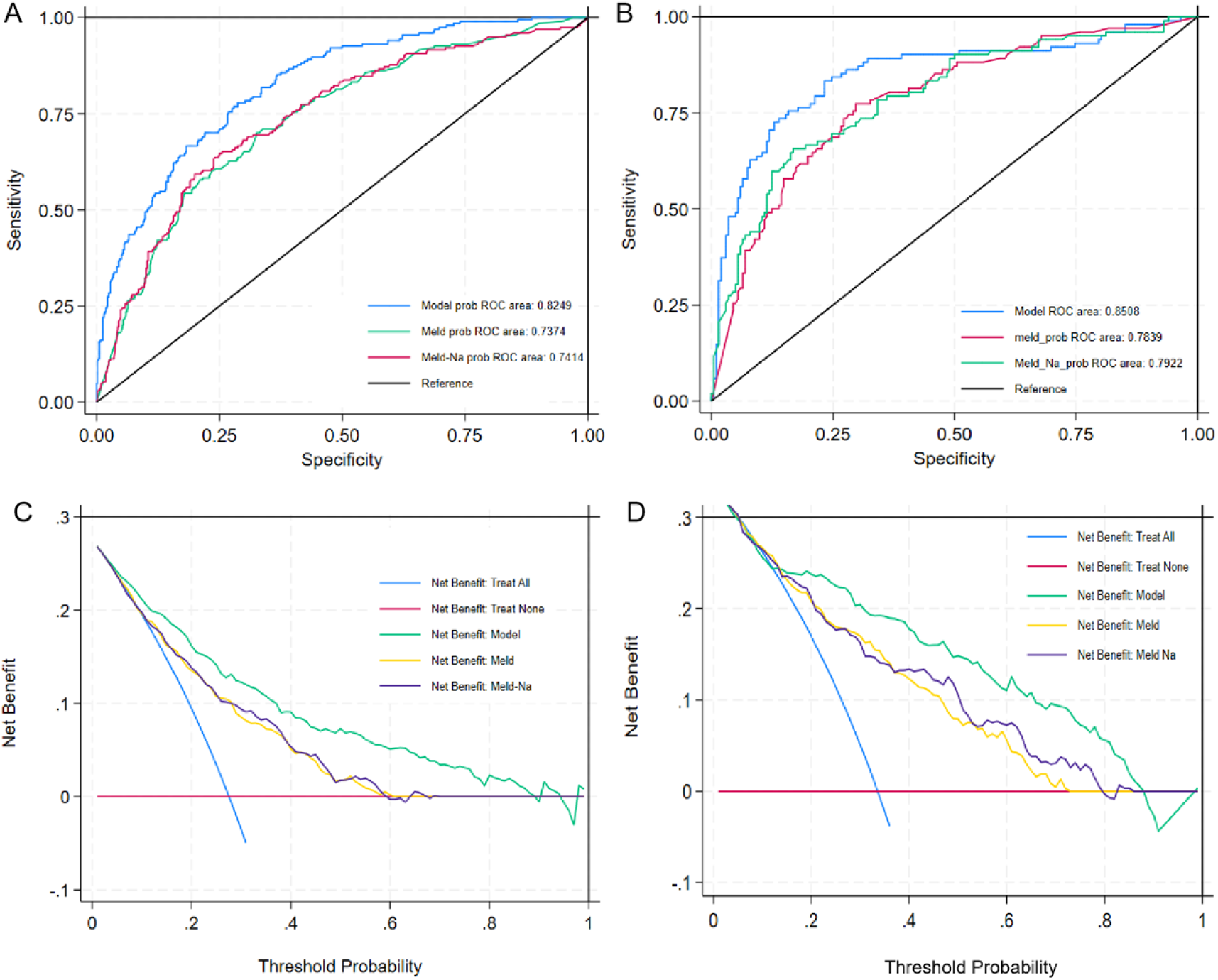
Clinical utility and predictive performance evaluation of the short-term mortality model in severe cirrhosis. (A) Receiver operating characteristic (ROC) curve comparisons of multiple predictive models in the derivation cohort. The proposed model (red curve) achieved superior discriminative performance (AUC = 0.825, 95% CI: 0.793– 0.857) compared to conventional scoring systems. (B) External validation of model performance in an independent cohort, demonstrating maintained predictive accuracy AUC = 0.851 (95% CI: 0.800–0.901). (C) Decision curve analysis (DCA) in the derivation cohort showing enhanced net benefit of the proposed model (blue curve) across clinically relevant threshold probabilities (20–80%), outperforming existing models. (D) Validation cohort DCA confirming consistent clinical utility of the novel model (blue curve) compared to standard approaches.

#### Clinical Utility Assessment

Decision curve analysis (DCA) evaluating threshold probabilities from 20% to 80% demonstrated the model’s superior clinical utility. Across this clinically actionable range, the nomogram provided higher net benefit than both “treat-all” and “treat-none” strategies, outperforming MELD and MELD-Na scores in derivation and validation cohorts (Figures 4C and 4D). This translates to enhanced capacity for avoiding overtreatment in low-risk patients while appropriately prioritizing high-risk individuals requiring intervention.

### 3.4 Short-term Prognostic Evaluation Using Cox Regression

Univariate analysis identified 13 variables significantly associated with survival time (P<0.05), including laboratory markers (AST, INR, creatinine, hemoglobin, platelets, BUN, WBC, albumin, sodium, total bilirubin, ALP, RBC), comorbidities (renal disease, peptic ulcer disease, metastatic solid tumors), and age. Multivariable Cox regression (Table 4; Figure 5A) revealed independent predictors across five clinical domains: (1) Hematologic: elevated red blood cell count (adjusted hazard ratio [aHR] 0.791, 95% CI 0.626-1.000 per 10⁶/μL increase) and platelet count (aHR 0.997, 95% CI 0.995-0.998 per ×10³/μL) conferred protective effects; (2) Hepatic: each 1-U/L increase in AST (aHR 1.000, 95% CI 1.000-1.001) and 1-mg/dL rise in total bilirubin (aHR 1.031, 95% CI 1.014-1.048) amplified mortality risk; (3) Coagulation: INR elevation (aHR 1.078, 95% CI 1.021-1.139 per unit); (4) Renal: 1-mg/dL creatinine increase (aHR 1.112, 95% CI 1.065-1.161); (5) Comorbidities: metastatic solid tumors doubled mortality risk (aHR 2.023, 95% CI 1.151-3.553; P<0.001), whereas peptic ulcer disease showed paradoxical protection (aHR 0.467, 95% CI 0.229-0.952; P=0.036). Notably, every 10⁶/μL RBC increase reduced mortality risk by 20.9%, establishing hematologic stability as a key survival determinant.Bootstrap validation revealed distinct prognostic patterns across covariates. Age, AST, INR, creatinine, platelet count, WBC, and total bilirubin demonstrated narrow 95% confidence intervals (CIs) excluding the null hazard ratio (HR=1), indicating statistically significant and precisely estimated effects on short-term prognosis. Conversely, while peptic ulcer disease and metastatic solid tumors showed significant associations (CIs excluding HR=1), their wide confidence intervals reflected clinically relevant but imprecisely quantified prognostic effects. Red blood cell count (RBC) exhibited a 95% CI spanning HR=1, confirming non-significant prognostic value. Critically, bootstrap-derived HR estimates demonstrated full concordance with original Cox regression results (mean absolute difference <0.05) (Supplementary Table 1), validating model stability and supporting clinical utility for mortality risk prediction.

**Table4.**
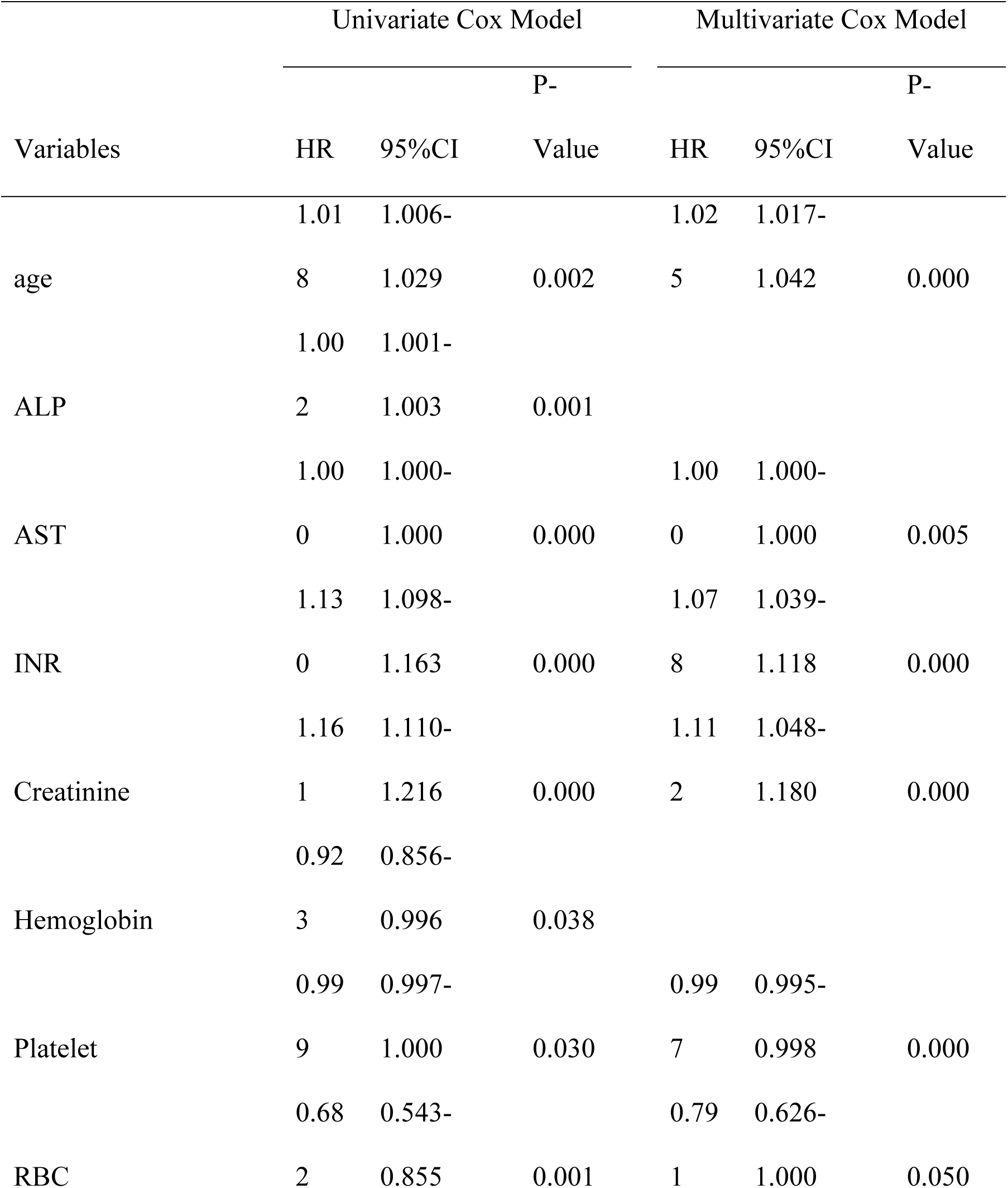
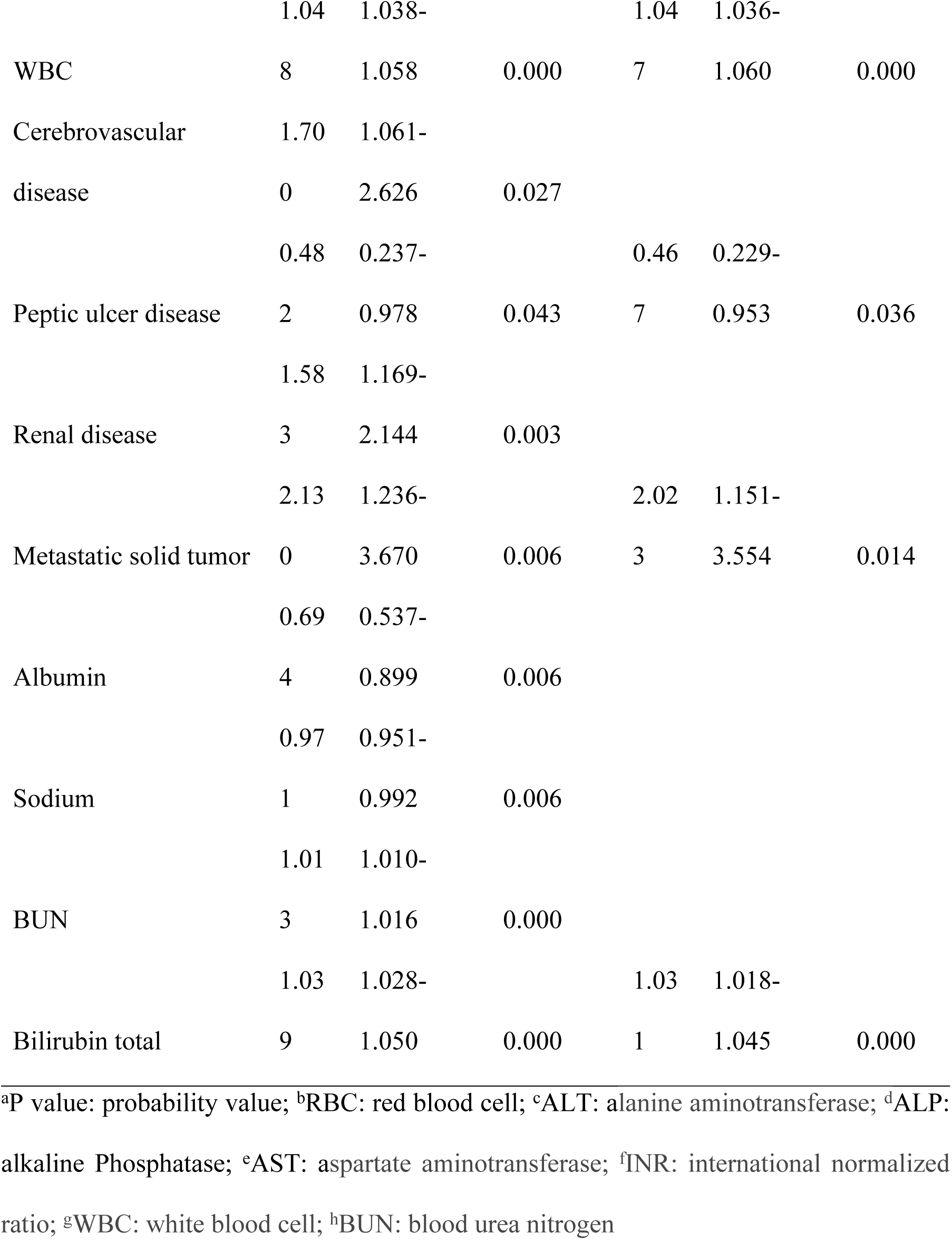
Univariate and multivariate Cox regression analyses.

**Figure 5.**
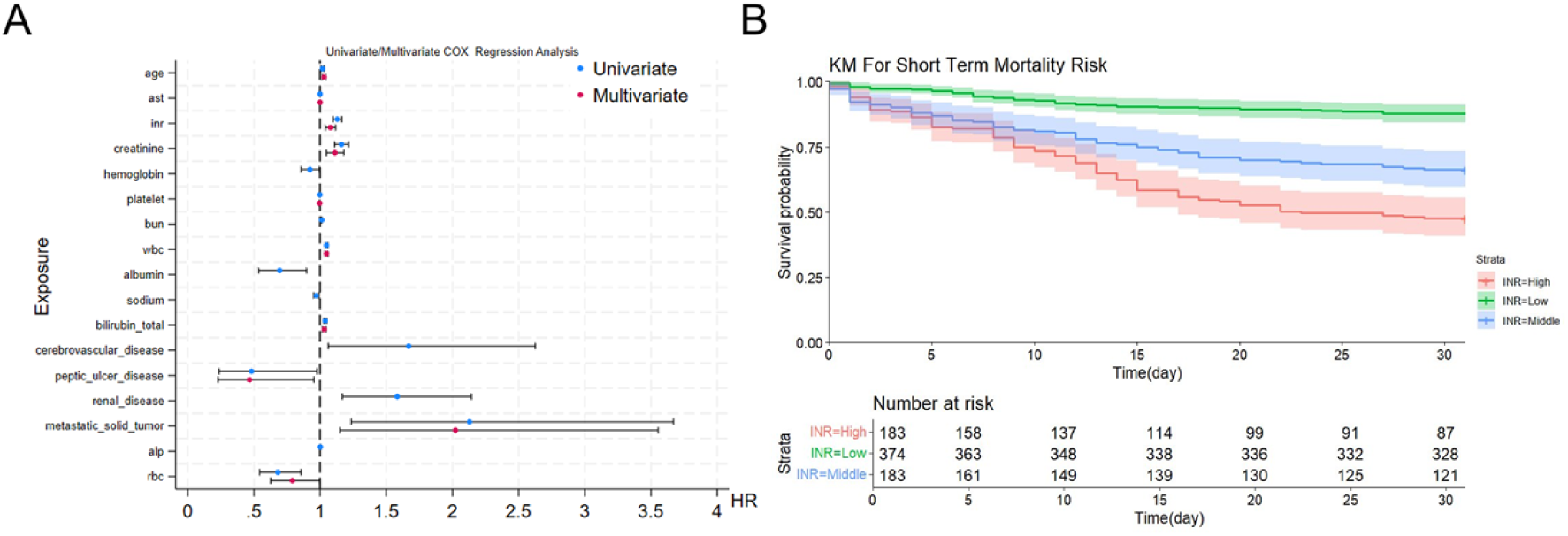
Prognostic analysis of severe cirrhosis patients. (A) Forest plot of risk factors associated with 30-day mortality derived from univariate and multivariate Cox regression analyses. Significant predictors (*p* < 0.05) are highlighted, with hazard ratios (HR) and 95% confidence intervals (CI) shown for key variables. (B) Kaplan-Meier survival curves comparing short-term outcomes across distinct INR strata (<1.7, 1.7–2.7, >2.7). Log-rank tests confirmed significant survival differences between groups (*p* < 0.001), with INR >2.7 demonstrating the poorest prognosis.

### 3.5 INR Stratification Analysis

Using optimal prognostic cutoffs derived from X-tile analysis, we stratified patients into three clinically distinct INR risk categories: low-risk (<1.7, reference), intermediate-risk (1.7–2.7), and high-risk (>2.7). Kaplan-Meier survival curves (Figure 5B) demonstrated significant intergroup disparities (log-rank P<0.001), with mortality risk escalating proportionally to INR levels: high-risk (30-day survival 47.4%) > intermediate-risk (66.1%) > low-risk (87.7%). In our validation, such hierarchical differences are also consistent(Supplementary Figure1).This graded association underscores INR as a continuous risk modulator in cirrhosis prognosis, providing actionable thresholds for intensity-adjusted monitoring.Internal validation independently confirmed these associations.

**Supplementary Figure 1. Kaplan-Meier survival curves comparing short-term outcomes across distinct INR strata (<1.7, 1.7–2.7, >2.7) in validation**. Log-rank tests confirmed significant survival differences between groups (*p* < 0.001), with INR >2.7 demonstrating the poorest prognosis.

## 4 Discussion

### 4.1 Key Findings and Model Development

This study presents the first comprehensive predictive model for short-term mortality risk in patients with severe cirrhosis, incorporating an analysis of short-term prognosis and evaluating the prognostic significance of the international normalized ratio (INR). Our findings demonstrate that the prediction model exhibits excellent sensitivity and specificity, with short-term prognosis being significantly associated with multiple factors including age, aspartate aminotransferase levels, INR, creatinine, platelet count, white blood cell count, total bilirubin levels, red blood cell count, and the presence of comorbidities such as peptic ulcer disease and metastatic solid tumors. Notably, we observed substantial variations in survival prognosis among patients with different INR levels.

### 4.2 Multidimensional Predictors of Short-Term Mortality in Severe Cirrhosis

The high short-term mortality risk and poor survival prognosis in severe cirrhosis patients underscore the clinical importance of accurate risk prediction. Our analysis revealed that each additional year of age increased short-term mortality risk by 1.04-fold, consistent with existing literature documenting the critical impact of age-related physiological changes on cirrhosis outcomes. These changes include deterioration in circulatory dynamics, organ function, and immune competence, all of which significantly influence the development of cirrhosis-related complications[19]. The strong association between age and prognosis in our study confirms its value as a reliable predictor of short-term mortality in severe cirrhosis.

Renal function emerged as another crucial prognostic factor, with elevated creatinine levels demonstrating significant predictive value. While the precise mechanisms linking cirrhosis to renal impairment remain incompletely understood, our findings support existing research identifying multiple contributing factors: hypovolemia (50% of cases), intrinsic structural kidney damage (30%), hepatorenal syndrome (15-20%), and postrenal obstruction (<1%)[20, 21]. These diverse pathways collectively impair renal excretory function, leading to creatinine elevation and corresponding increases in mortality risk.

Hematological parameters provided additional prognostic insights. Thrombocytopenia showed an inverse relationship with survival, potentially attributable to both decreased platelet production (mediated through reduced thrombopoietin activity in fibrotic livers[22]) and increased platelet destruction (via hypersplenism and immune-mediated clearance[23, 24]). White blood cell count served as an independent predictor of mortality, reflecting the high prevalence (24-29%) of bacterial infections in advanced cirrhosis and their substantial contribution to poor outcomes[25, 26]. The inclusion of this parameter addresses a notable gap in previous prediction models that often overlooked infection-related mortality.

Total bilirubin levels demonstrated significant prognostic value, with elevations resulting from both decreased hepatic clearance (direct bilirubin) and increased hemolysis (indirect bilirubin)[27–29]. Interestingly, our model identified a potentially protective association between peptic ulcer disease and reduced mortality, possibly reflecting the benefits of ulcer-related therapeutic interventions (e.g., proton pump inhibitors, antimicrobial prophylaxis) in mitigating bleeding risk and other complications[30, 31]. Conversely, metastatic solid tumors were associated with a threefold increase in mortality, consistent with their characteristic disease progression patterns and treatment resistance[32, 33].

The prognostic value of INR stratification represents another key finding, with higher levels correlating with poorer outcomes. This relationship likely reflects both impaired hepatic synthesis of coagulation factors[34, 35] and platelet-related abnormalities[24, 36]. Our results align with prior research demonstrating INR’s role in bleeding risk assessment, while also highlighting its broader utility in mortality prediction[37].

### 4.3 Methodological Innovations

This study is distinguished by three key advancements: the systematic integration of comorbidities, addressing oversights in previous models; the use of extreme-value laboratory analysis to capture critical physiological states more accurately than averaged measures; and the introduction of clinically actionable INR stratification, replacing traditional binary classifications. These innovations yielded a model demonstrating superior discrimination compared to existing scores while maintaining practicality through the use of routine parameters.

### 4.4 Study Limitations

Several constraints warrant consideration: the model’s derivation from the single-center MIMIC-IV ICU database limits generalizability to non-critical care settings; the exclusion of cases with >20% missing data (affecting 18% of screened patients) may introduce selection bias; and the use of conventional regression approaches, while ensuring clinical interpretability, may lack the precision of machine learning algorithms. These limitations collectively highlight the necessity for future multicenter validation studies incorporating advanced analytical methods.

## 5 Conclusions

This study establishes a clinically implementable prediction model for short-term mortality in severe cirrhosis, demonstrating superior performance to traditional prognostic scores through innovative parameter selection and INR stratification. While our findings provide critical insights into comorbidity-associated risk modulation and coagulation parameter interpretation, prospective validation across diverse care settings remains essential prior to clinical implementation. Future research directions should prioritize integration of dynamic laboratory trends and therapeutic response biomarkers to enhance predictive precision.

## Author contributions

Shun Zhang contributed to data extraction, analysis, and manuscript drafting. Rui Liu performed statistical analyses and created the tables. Zhengjie Li participated in data interpretation and prepared graphical illustrations. Tao Pan and Xudong Wen oversaw data validation, critically reviewed the manuscript, and provided substantive revisions. All authors approved the final version for submission.

## Acknowledgements

We acknowledge the MIMIC dataset for offering diverse data resources, allowing us to pursue comprehensive and profound research.

## Data Availability Statement

The datasets analyzed in this study were derived from the MIMIC-IV database (https://physionet.org/content/mimiciv/).

